# Direct Primary Care: Family Physician Perceptions of a Growing Model

**DOI:** 10.1101/2021.03.22.21254097

**Authors:** Gayle Brekke, Jarron M. Saint Onge, Kim S. Kimminau, Shellie Ellis

## Abstract

**Purpose:** Direct Primary Care (DPC) is a relatively new primary care practice model in which patients receive unlimited access to a defined set of primary care services in exchange for a monthly practice-specific membership fee. DPC is a bottom-up physician-driven approach that contrasts to typical top-down insurer-centric health care delivery reform efforts. The degree to which physicians are aware of this practice model and whether they believe it addresses two key challenges facing primary care, access and administrative burden, are unclear.

**Methods:** An online survey distributed in July 2017 gauged family physicians’ awareness of DPC and views about the model.

**Results:** Most respondents (85%) had heard of DPC and eight percent practiced in a DPC model at the time of the survey. In general, respondents reported that DPC can offer positive outcomes through lower administrative burden for physicians, improved doctor-patient relationships, and better access. Respondents also suggested DPC may result in improved patient health outcomes and lower overall healthcare spending. Respondents’ concerns included inappropriateness of the model for vulnerable populations and physician shortages. Survey responses differed depending on whether the respondent practiced in a DPC model; DPC physicians had a more favorable view of the model and were focused on benefits to patients rather than benefits to physicians.

**Conclusions:** While some perceive challenges of DPC, others think that this model may benefit both patients and physicians.

## INTRODUCTION

Despite widespread agreement that the availability of robust primary care is critical to the health of patients and the performance of the health care system overall,^1-3^ primary care faces several difficult challenges, including a lack of adequate access for many patients,^4-6^ and high levels of administrative burden leading to both physician burnout and career dissatisfaction.^7-12^ While alternative models of primary care delivery, such as Direct Primary Care (DPC), offer the potential to mitigate these challenges, less is known about physicians’ knowledge and perspectives on the relevance of new approaches to care. Accordingly, this study uses a sample of family physicians to explore their understanding and interest in adopting a DPC approach and how it varies by their current practice model.

DPC is a primary care delivery model characterized by enhanced access to care;^13^ DPC patient panels are roughly one-third to one-fourth the size of typical PCP patient panels and DPC physicians spend a smaller portion of their time on administrative tasks.^14,15^ In a policy statement supporting DPC in December 2017, the American Academy of Family Physicians (AAFP) said they found DPC to be consistent with the Academy’s advocacy to protect and enhance “the intrinsic power of the relationship between a patient and his/her family physician to improve health outcomes and lower overall health care costs.” A body of literature connects various aspects of the doctor-patient relationship and trust with patient health outcomes such as use of preventive services, treatment adherence, reduced ED visits and hospitalizations, and lower expenditures.^16-19^

Several articles and commentaries suggest that DPC practices have benefits specific to physicians, as well as to patients. For example, emerging research shows the potential for improved physician satisfaction through patient engagement and shared decision making, increased time to develop personal relationships and improvement in the quality of care.^20-23^ Additionally, DPC models have the potential to reduce administrative burden for physicians. DPC is a decentralized, physician-driven model of primary care delivery compared to other more common delivery reform efforts that tend to be more centralized and bureaucratic. For example, delivery reform efforts such as patient-centered medical homes and accountable care organizations are characterized by guidelines and oversight, which may serve patients, but may also be associated with high administrative burden among physicians due to their numerous documentation and reporting requirements.^8,9,24^ In contrast, there is no national DPC organization to define requirements or practice rules, and there are no documentation or reporting requirements associated with the DPC model, which may reduce physician administrative burden but may also have implications for practice standards.

This study gauges the awareness of DPC among a sample of family physicians. Specifically, the objective is to understand whether physicians are aware of DPC and if they view DPC as a model with potential benefits. We explore physician perceptions of DPC patient care and their understanding of the model. Next, we aim to understand physician perceptions about the model’s potential to increase professional satisfaction. Finally, we compare responses by the respondent’s practice model (DPC compared to non-DPC) to describe characteristics germane to both groups.

## NEW CONTRIBUTION

There remains limited visibility of the DPC model and it is unclear how physicians understand or interpret the potential advantages or challenges associated with this approach. Physician perspectives about DPC are important for several reasons. First, physician-level knowledge about the structure and definitions of a DPC model will provide information about whether physicians accurately understand the model. Second, if uptake and/or modifications to the DPC model are to increase, it remains important to understand how family medicine physicians perceive advantages and disadvantages of the model. Questions about whether physicians perceive that DPC might improve career satisfaction or improve patient access to primary care present an opportunity to assess the potential growth of the DPC model in the US. Third, research consistently shows that administrative burden (leading to physician burnout and career dissatisfaction) and lack of adequate access to primary care for some patients remain key problems in primary care.^4-12^ Identifying the differences in perceptions among DPC compared to non-DPC physicians will highlight whether those currently in the model exhibit more or less favorable perceptions of a DPC approach.

## METHODS

We developed a survey that was distributed online by the American Academy of Family Physicians (AAFP) during the week of July 17, 2017. Participants were self-selected through their participation in the Member Insight Exchange (MIE), the marketing research online community of the AAFP, which had a total of 672 members. Results of MIE surveys, while not fully generalizable, provide a snapshot of views and practice patterns of AAFP members.^25,26^The Institutional Review Board of the University of Kansas Medical Center determined the study is exempt from human subjects protections review. Most survey questions had a small number of fixed responses and respondents had an opportunity to submit written responses to open-ended questions. Items elicited physicians’ perceptions about 1) patients’ understanding of DPC, 2) the financial sustainability of DPC practice, 3) patient health outcomes in DPC, and 4) quality of care in DPC.

Respondents were first asked whether they have heard of the DPC model and whether they currently practice in this model; they were then asked to choose the best definition of DPC from a short list in which generally accepted definitions of DPC, patient centered medical homes (PCMHs), and other primary care delivery models were provided as options.

Second, we used an AAFP-endorsed definition to assess knowledge and beliefs about the DPC model. AAFP defines DPC as a model in which physicians do not accept payments from insurance companies or other third-party payers, but rather, patients pay a monthly membership fee ranging from $50 to $150 for a defined set of primary care services for no extra charge, and low-cost prescriptions and other services.^13^ Survey respondents were asked to indicate their agreement on a Likert-type scale with each of several statements about the model. Statement #1: Patient confusion. Statement #2: Not financially sustainable. Statement #3: Increases physician shortage. Statement #4: Only benefits healthy & wealthy. Statement #5: Lack of health improvement. Statement #6: Low quality. Statements were worded negatively with agreement indicating negative perceptions of DPC. See Figure 1 for complete text of the statements.

**Figure 1.**
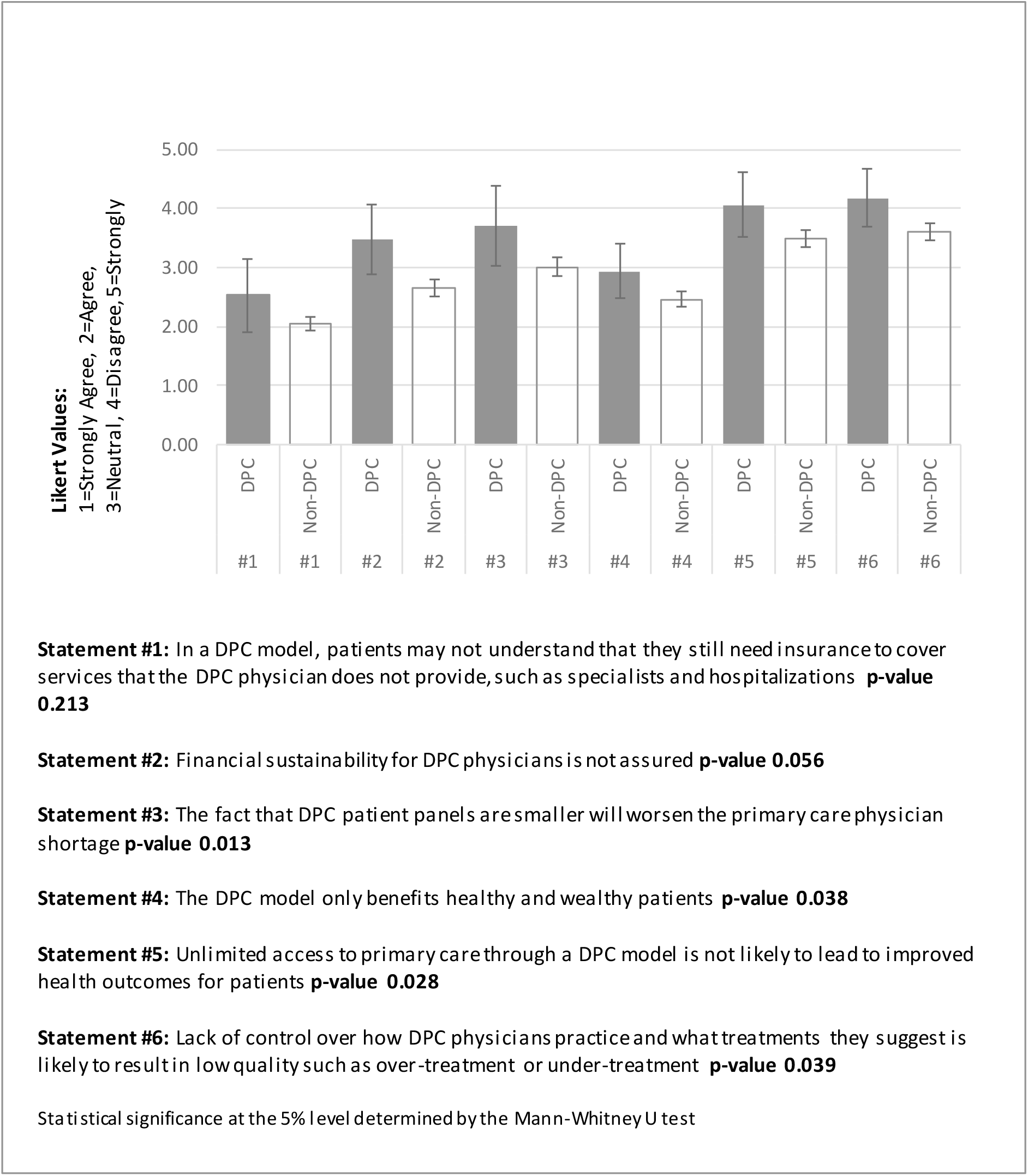
Level of Agreement with Statements about Direct Primary Care, by Practice Model 95% Confidence Level

Next, respondents were asked to rank the top three benefits of the model, selecting from a list of five choices. Respondents could select “Other” and enter a written response. The topics included administrative burden, time with patients, spending on downstream care, and physician responsiveness to patient needs and preferences.

Results were summarized and compared by practice model (DPC or not), using the Mann Whitney U test. Complete case analysis was performed. Fifteen surveys with missing data on either the outcome or the belief statements were excluded.

## RESULTS

There were 225 complete responses to this survey for a response rate of 33%. Respondents were similar to AAFP membership by sex, years since completed residency and practice ownership model. Table 1 represents distributions of survey respondents, MIE members and AAFP members, however, variables of interest to this study are not available in the broader data.

**Table 1.** Characteristics of Survey Respondents, Member Insight Exchange and AAFP Active Members (2017 AAFP data)

|  | **AAFP** | **Member Insight**<br>**Exchange** | **Survey**<br>**Respondents** |
| --- | --- | --- | --- |
| **Sex** |  |  |  |
| Male | 58% | 50% | 55% |
| Female | 42 | 50 | 45 |
| **Years Since Completed Residency** |  |  |  |
| 7 or fewer | 25 | 32 | 21 |
| 8 to 14 | 22 | 23 | 22 |
| 15 to 21 | 21 | 27 | 21 |
| 22 or more | 32 | 19 | 37 |
| **Ownership Model** |  |  |  |
| Sole owner | 13 | 13 | 11 |
| Partial owner | 17 | 14 | 12 |
| 100% employed | 66 | 69 | 74 |
| Not applicable / not in clinical<br>practice | 3 | 3 | 2 |
| **Member Count** | 68,300 | 672 | 225 |
| No variables are significantly different between AAFP Members and Survey Respondents |  |  |  |

Most respondents (85%) were familiar with DPC, with eight percent of the sample reporting practicing in a DPC model; a majority (79%) selected the AAFP-endorsed definition of DPC, 13% responded “Don’t know” and eight percent selected a definition that aligns with PCMH rather than DPC.

Respondents indicated agreement with statements that DPC has benefits for patients. For example, a minority expressed concern about quality in DPC, with 29 (13%) perceiving that DPC practice will result in quality problems such as over-treatment and under-treatment.

Similarly, 43 (19%) perceived that DPC patients will not experience improved health outcomes. There was, however, concern about the applicability of DPC to all patients. While the survey did not directly address low income or vulnerable patients, two respondents expressed this concern in the open-ended responses. One physician stated, “This model may leave undue burden on patients and it risks losing the primary care safety net for the very poor and underserved.”

For all statements included in Figure 1 agreement was greater among non-DPC than DPC family physicians. For four of the six belief statements, the difference in responses by the physician’s current practice model was statistically significant. Compared to non-DPC physicians, DPC physicians were less concerned about the primary care physician shortage (p=0.013), less concerned about the potential for over-treatment and under-treatment in DPC (p=0.039), more certain that DPC benefits more than just healthy and wealthy patients (p=0.038), and more certain that DPC will lead to improved patient health outcomes (p=0.028).

Figure 2 presents the respondent rankings of the top-three most important benefits of DPC. Overall, respondents ranked the most important benefits about DPC as 1) lower administrative burden, 2) improved patient health outcomes, and 3) lower health care expenditures. Forty-eight percent of respondents selected lower administrative burden as the most important benefit and 84% selected it as one of the top 3 benefits of DPC.

**Figure 2.**
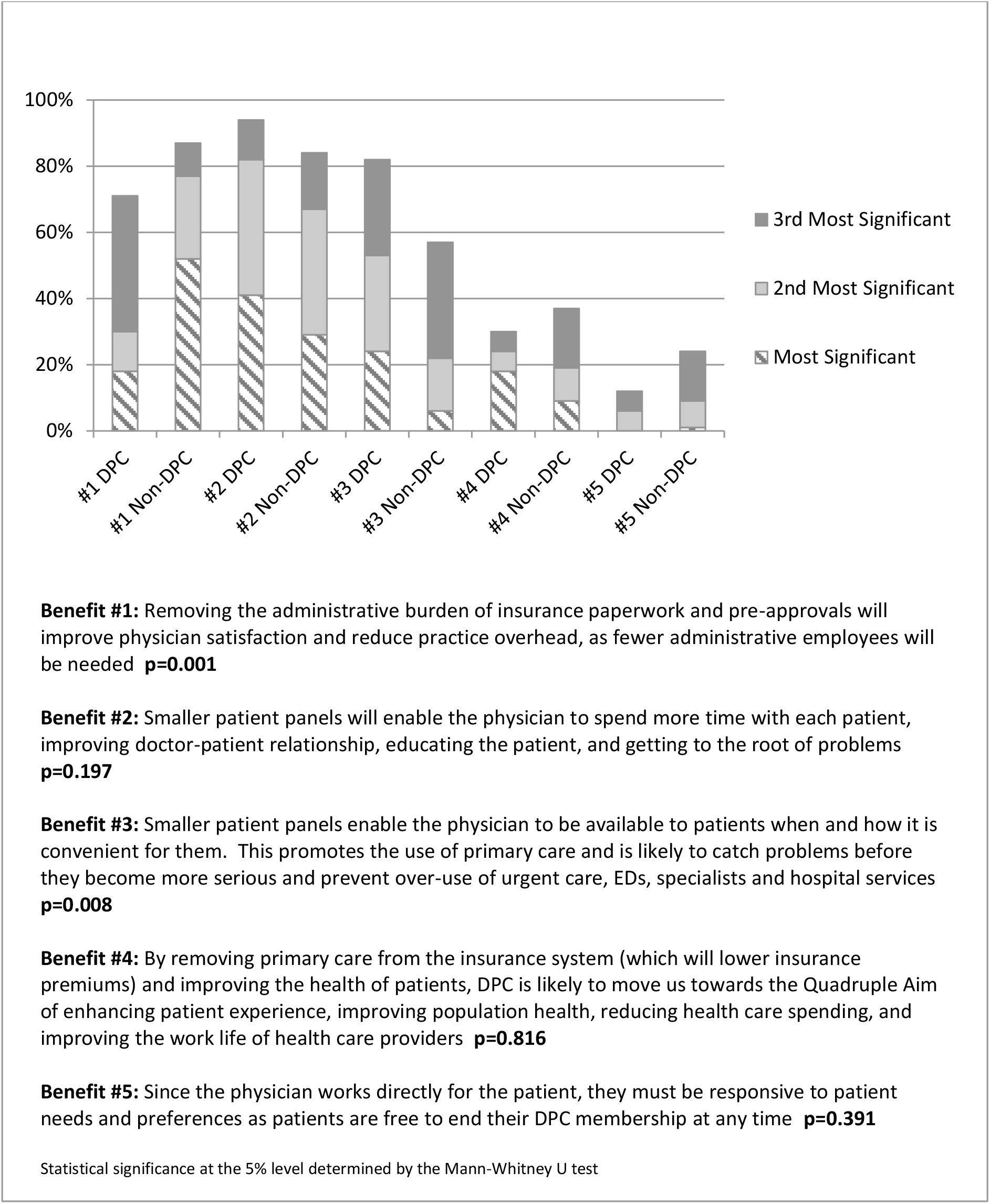
Rankings of DPC Benefits by Practice Model

Increased time with patients to improve the doctor-patient relationship and to educate the patient and get to the root of problems was selected by 68 respondents (30%) as the most important benefit and by 189 (84%) as one of the top 3 benefits of DPC. Better availability of care which prevents overuse of downstream services was only selected as the most important benefit by 18 respondents (8%), 133 (59%) selected it as one of the top benefits.

Rankings were statistically significant by practice model. Lower administrative burden was selected as the most important benefit by half (51%) of non-DPC physicians, but only 18% of DPC physicians (p=0.001). Nearly 25% of DPC physicians selected better availability of care as the most important benefit of DPC; 6% of non-DPC physicians made the same selection (p=0.008). The three other benefits were not significantly different by practice model.

## DISCUSSION

The goal of this study was to compare familiarity and perceived benefits of a DPC model by DPC and non-DPC family practitioners to understand the potential for physician satisfaction. This is the first survey to assess family physicians’ views about Direct Primary Care informed by whether they practice in the model. Several commentaries and opinion pieces have been published on DPC^27-29^ but no studies distinguish between perspectives held by those practicing and those not practicing in a DPC model. In previous work, DPC either was not included in physician surveys, or was reported only in combination with other alternative payment models, which obscured model-specific physician views of DPC.^30^ The variation in perspectives may be relevant to how broadly suitable DPC is for various physicians and patient populations, and thus its growth potential and its applicability in various settings.^22,26^ By highlighting these differences, we aimed to understand the extent to which DPC models are understood and its perceived benefits to physicians and patients. Overall, we found that family medicine physicians who responded to this survey share a high level of familiarity with DPC, with 85% of respondents having heard of the model and 79% correctly identifying the overall goal of the model. Beginning with this highly knowledgeable group, we demonstrated fairly consistent perceptions of the benefits but with a few key distinctions by whether or not they practiced in a DPC model.

First, we found that family physicians broadly agree that DPC has the potential to improve patient health outcomes. More time with patients, improving the doctor-patient relationship and getting to the root of problems were tied as the most significant benefit of the model, while the statement that patient health outcomes won’t improve in DPC received high disagreement. This finding supports studies that show a positive association between improved doctor-patient relationships and improved patient health outcomes.^16,17,19^

Second, we found that family physicians generally agree that administrative burden is less in a DPC model, which will improve physician satisfaction. Improved physician satisfaction due to lower administrative burden and less paperwork was tied as the most significant benefit of the model. This finding supports what other studies have found regarding physician burnout and dissatisfaction resulting from high levels of administrative burden.^7-12^

Third, family medicine physicians think that DPC is beneficial for both patients and physicians; this, along with limited concerns about physician shortages and access for vulnerable patients, may suggest potential success of bottom-up physician-driven models more generally. In particular, respondents did not perceive that patient outcomes or quality would likely suffer without the additional documentation and reporting tasks that are required by typical top-down centralized health care delivery reform initiatives. Respondents expressed high disagreement to the statement that DPC will result in low quality such as over-treatment or under-treatment. This view is shared by physicians and scholars who say that primary care does not lend itself to meaningful performance measurement due to its complexity and uncertainty. More than a third of health concerns initially encountered in primary care do not lend themselves to a diagnosis, and about half are unlikely to result in a definite diagnosis that would trigger a standard care pathway.^31^ Complex interactions and interdependencies emerge in primary care due to the many hundreds of decisions that must be made each day for patients with unique clinical concerns and personal circumstances.^32^ Typical performance measures cannot address the breadth and depth of comprehensive primary care delivery; simply measuring individual elements of care is an inadequate reflection of the value of primary care.^33^

Finally, this study adds to the literature by showing significant variation in the perspectives of family medicine physicians who practice in an insurance-based model and those who practice in a DPC model. DPC physicians are likely to be highly select in this sample, with both experience and perceptions of the model. DPC physicians expressed more disagreement with all of the statements, and the differences were statistically significant for four of the six statements. As all statements were negatively worded, this suggests that DPC physicians were more confident about the DPC model than their non-DPC colleagues. In addition, the finding that DPC physicians were much less focused on the perceived benefit of reduced administrative burden than the non-DPC family physicians may indicate that administrative burden is less a part of their day to day practice of medicine than it is for non-DPC physicians.

The study has several limitations. First, the survey respondents are likely to be a highly select group. While we found very similar demographic profiles to the overall membership, the results may not be generalizable to all family physicians or to the AAFP membership. AAFP’s Member Insight Exchange is a marketing platform in which respondents self-select to receive surveys and may choose to answer or ignore any survey provided; these results are likely not representative of overall family physicians’ views of DPC. Physicians who are familiar with DPC or who have a strong opinion about DPC may have been more likely to respond to the survey than physicians who are unfamiliar with the model or who have not yet formed an opinion. For example, these results differ from results of AAFP’s 2017 Practice Profile Survey, which showed that one-third of family physicians (33%) were unfamiliar with the DPC model, a small portion (3%) practiced in a DPC model, and a small portion (1%) were in the process of transitioning their practice to a DPC model.^26^ Given the limited scope of this survey and that it was available to a small subset of AAFP members, further research is needed to gauge family physician views of DPC more broadly and to improve the generalizability of these findings. However, this study offers insights beyond generalizability.

First, it allows us to understand perceptions from a group that both understands the model and has ostensibly strong feelings (both positive and negative) about the model. While the study is not comprehensive, these respondents are potentially the individuals more likely to propose or resist a move to a DPC model.

Second, this study was focused on the primary perceived benefits of a DPC model. Clearly, there are also strong challenges associated with this model that need to be addressed in future studies. Additionally, our list of benefits was not comprehensive, but rather, was consistent with previous literature’s suggested benefits.^3,16,18^ By focusing on these perceptions, we determined what areas and topics are agreed to by both DPC and non-DPC physicians.

This study revealed variation in perspectives about DPC between family physicians who practice in a DPC model and those who do not. While the survey does not allow us to determine why perspectives differ by practice model, there are several possible explanations. Non-DPC physicians may have limited familiarity with how DPC works in practice, and their lack of familiarity may cause them to be cautious about the model’s benefits. Conversely, DPC physicians and particularly those who responded to the survey were more likely to advocate the benefits of the model. Both perspectives should be approached with caution. Also, it is possible that the patient panels of non-DPC physicians are different from those of DPC physicians in ways that would affect the perceived success of the model for those physicians.

Observed differences in responses may only reflect differences in perceptions and may not be indicative of how care is objectively delivered in a DPC model compared to a traditional delivery model. It is also possible that variations in physician perspectives by model reflect real differences in patient outcomes and physician satisfaction due to the model. Additional studies are needed to test these conjectures.

## Conclusions

In general, respondents perceive that Direct Primary Care offers more time with patients and improved doctor-patient relationships, which may lead to improved patient outcomes and lower health care spending. Despite concerns about physician shortages and access for vulnerable patients, respondents perceived the potential of DPC to reduce administrative burden, resulting in greater physician satisfaction and lower practice costs. They believe that these patient and physician benefits are available in DPC without negative impacts to quality such as over-treatment and under-treatment.

Compared to non-DPC physicians, views of family physicians practicing in a DPC model were found to differ in several areas. DPC physicians exhibited statistically significantly higher disagreement with statements that DPC only benefits the healthy and wealthy, it will worsen the PCP shortage, it will not lead to improved patient health outcomes, and quality will be poor. Additionally, DPC physicians focused more on perceived benefit to patients (enhanced access to primary care, resulting in improved patient health outcomes) while non-DPC physicians focused more on perceived benefits to physicians (less administrative burden and insurance paperwork, improved physician satisfaction). Our survey is the first to compare the views of physicians by practice model but further research is needed to understand the nature and causes of these differences.

These results are important initial findings about a growing model of primary care delivery. Additional research is needed to determine whether DPC delivers the kinds of benefits that are integral to the rationale and structure of the model, and to illuminate DPC’s potential to address access, cost and workforce-related problems facing primary care and family medicine today.

## Data Availability

I do not wish to make data publicly available.

## Acknowledgements

This survey was made possible with assistance from the American Academy of Family Physicians National Research Network

